# Selective vulnerability of human basal forebrain subsystems in ageing and neuropsychiatric disorders

**DOI:** 10.64898/2026.10.13.26365385

**Authors:** Tamir Eisenstein, Michele TM Hu, Johannes C. Klein

**Affiliations:** Oxford Centre for Integrative Neuroimaging, FMRIB, Nuffield Department of Clinical Neurosciences, University of Oxford, Oxford, UK; Oxford Parkinson’s Disease Centre, Division of Neurology, Nuffield Department of Clinical Neurosciences, University of Oxford, Oxford, UK

**Keywords:** Basal forebrain, Ageing, Dementia, Structural MRI, Lifespan, Neurodegeneration

## Abstract

**INTRODUCTION:** The basal forebrain (BF) provides the principal cholinergic innervation to the cortex and the hippocampus and is among the earliest regions affected by neurodegenerative processes. Yet, whether its subnuclei differ in how their structural integrity changes with age and in their vulnerability to neurological and psychiatric disorders is unresolved.

**METHODS:** Here, in 81,870 UK Biobank participants aged 45–87 years, we built age- and sex-specific normative models of grey-matter volume in the nucleus basalis of Meynert (Ch4) and the septal/diagonal-band (Ch1–3) compartments of the BF and derived individual deviation from these norms. We then tested the selectivity of their normative deviation profile across cross-sectional and longitudinal ageing, manifest neurological and psychiatric disorders, pre-diagnostic neurodegeneration, prospective disease incidence, and neuropsychiatric polygenic liability.

**RESULTS:** The two compartments followed distinguishable age-related profiles across mid-to-late adulthood, which persisted when examined using longitudinal data. Deviation from norms was generally concentrated in Ch4 across neurological diseases and, in almost no condition, greater in the Ch1-3. The Ch4 deficit was further detectable years before diagnosis, was not substantially explained by common inherited genetic risk, and was linked with incident dementia and Parkinson’s disease, whereas Ch1–3 carried no independent prospective structural vulnerability.

**DISCUSSION:** These findings identify an anatomically specific age-related profile of the human BF, with structural vulnerability manifest and pre-diagnostic neurological disorders, which is not substantially explained by common inherited risk.

## Introduction

The basal forebrain (BF) is the main source of acetylcholine (ACh) to the cerebral cortex, amygdala, and hippocampus, playing key roles in physiological, cognitive, and behavioural functions^1,2^. The neurons of the BF are organised into several cell groups of which the Ch1 is associated with cells in the medial septal nucleus (MSN), the Ch2-3 with the vertical and horizontal limbs of the diagonal band of Broca, and the Ch4 with the nucleus basalis of Meynert (NbM). These groups differ in their projection targets, with Ch1–2 providing the main cholinergic innervation to the hippocampus, Ch3 to the olfactory bulb, and Ch4 to the entire neocortex and amygdala^3^.

BF neurons are among the cell types most vulnerable to neurodegenerative processes, and their degeneration is an early event in both Alzheimer’s disease (AD) and Lewy body disorders (LBD) including Parkinson’s disease (PD), PD dementia, and dementia with Lewy bodies (DLB)^4–6^. The Ch4 in particular has been repeatedly implicated as an early site of vulnerability across neurodegenerative diseases. In AD, early post-mortem work reported profound (>75%) neuronal loss in Ch4, while later stereological analyses indicate that neuronal shrinkage and marker downregulation contribute to this^7,8^, while in vivo MRI studies showed Ch4 volume loss already at prodromal stages^9,10^. In LBD, Ch4 degeneration and cortical cholinergic denervation have been related to, and shown to predict, cognitive decline and dementia^11–13^.

However, much less attention has been given to the medial septum/diagonal band Ch1-3 subsystem despite it providing the principal cholinergic innervation to the hippocampus—the structure most closely tied to the amnestic syndrome of AD. While previous works provided evidence that Ch4 and Ch1–3 may behave differently in AD and LBD^8,14–17^, whether the two compartments are indeed differentially affected across diseases, and if so, how, still remains to be established, with recent reviews explicitly highlighting the need for disease-related, subregional-resolved analysis and for a clearer temporal staging of BF vulnerability^16,18^. More fundamentally, whether the two compartments even age differently in health remains unknown at the population level. The sub-regional evidence on BF ageing comes from a few relatively small cross-sectional cohorts^19^, and no population-scale reference model has elaborated the age-related profiles of Ch4 and Ch1–3. Furthermore, it remains unclear whether BF vulnerability is specific to neurodegenerative aetiology or reflects a broader susceptibility shared across neuropsychiatric disorders^20^.

Addressing these questions requires a framework that reflects vulnerability at the individual level, expressing each individual’s departure from expected norms at a given age and sex. Normative modelling offers both, by characterising the population distribution of a metric and then yielding an individual deviation score of departure from the expected value ^21–23^. Below-expected values can then be addressed as vulnerability, while above-expected values as resistance. Although such approaches have been applied to large-scale imaging cohorts, including UK Biobank (UKB)^21,24,25^, they have not addressed the BF, let alone its subnuclei. Given that the neurodegenerative process has been known to begin years before diagnosis^26^, individual-level markers of vulnerability ahead of clinical presentation are increasingly sought—to stratify risk in target populations, enrich prodromal cohorts for research and clinical trials, and characterise early stages of disease to aid mechanistic understanding.

Here we constructed age- and sex-related normative models of grey-matter volume in the Ch4/NbM and the medial Ch1–3/septal-diagonal compartments of the BF in the UK Biobank cohort and used them to address four questions: do the two compartments follow distinguishable age-related profiles across mid-to-late adulthood, establishing them as dissociable structures rather than a single, uniformly susceptible system? Is deviation from normative profiles differentially associated with major manifest neurological and psychiatric disease across the two compartments—and does the hippocampus-projecting Ch1–3 compartment feature in dementia as its connectivity would predict? Is such normative deviation detectable before diagnosis, and whether baseline deviation is associated with incident dementia an PD? And to what extent it tracks common genetic liability to these neuropsychiatric conditions?

## Methods

### Study design and participants

We used data from 81,870 UK Biobank participants aged 66.6±7.91 (range 45-87, 53% females) with available structural imaging data. The study followed the STROBE (Strengthening the Reporting of Observational Studies in Epidemiology) reporting guideline. Written informed consent of all participants was obtained by UK Biobank. Permission to use the UKB Resource was obtained as part of Data Access Application 95506. All participants provided written informed consent, and the study was approved by the National Information Governance Board for Health and Social Care and the National Health Service North-West Multicentre Research Ethics Committee (21/NW/0157, see http://www.ukbiobank.ac.uk/ethics/).

### Magnetic resonance imaging (MRI) and basal forebrain GM volume extraction

The acquisition protocol of the UK Biobank 3-Tesla T1-weighted structural imaging can be found here: https://biobank.ndph.ox.ac.uk/showcase/showcase/docs/brain_mri.pdf. The detailed preprocessing pipeline of the UK biobank imaging data has been previously described^27^. Images were processed with the UK Biobank brain imaging pipeline^28^, implemented primarily in FSL. In brief, T1w structural images were defaced, reduced in field of view, gradient distortion corrected, brain extracted using Brain Extraction Tool (BET)^29^, registered linearly and non-linearly to standard MNI152 space^30–32^, and segmented into tissue types (using FMRIB’s Automated Segmentation Tool (FAST)^33^, yielding partial volume estimate maps for grey matter, white matter and cerebrospinal fluid. Voxel-based morphometry was then performed on the native-space grey matter partial volume maps^34^. Each map was registered to a grey matter template in MNI152 space, and multiplied voxelwise by the Jacobian determinant of the non-linear component of the resulting warp field, producing modulated grey matter maps in which voxel intensities index grey matter volume. No spatial smoothing was applied prior to region-of-interest extraction, in order to preserve signal at the boundaries of the small structures examined here. Then, the GM volumes of the right and left Ch1-3 and Ch4 were extracted using masks from the probabilistic cytoarchitectonic map of the Basal Forebrain (v4.2)^35,36^, with a threshold of 50% following previous works and recommendations^37,38^ (**Figure 1**). The extracted BF metrics from both baseline and follow-up scans (see below) were then deconfounded for potential confound variables using linear mixed modelling (*lme4* package in R) including motion, scanner/table-position variables, T1-derived volumetric scaling factor, as well as UK biobank assessment centre and participant ID as random intercepts. For each imaging confound measure, values more extreme than 15 median absolute deviations (MAD) were excluded as potential outliers as previously suggested, as some confounders may have extremely non-Gaussian underlying distributions^39^. Similarly, participants with Ch1-3 or Ch4 values more extreme than 6 MAD were also excluded^27,39^.

**Figure 1.**
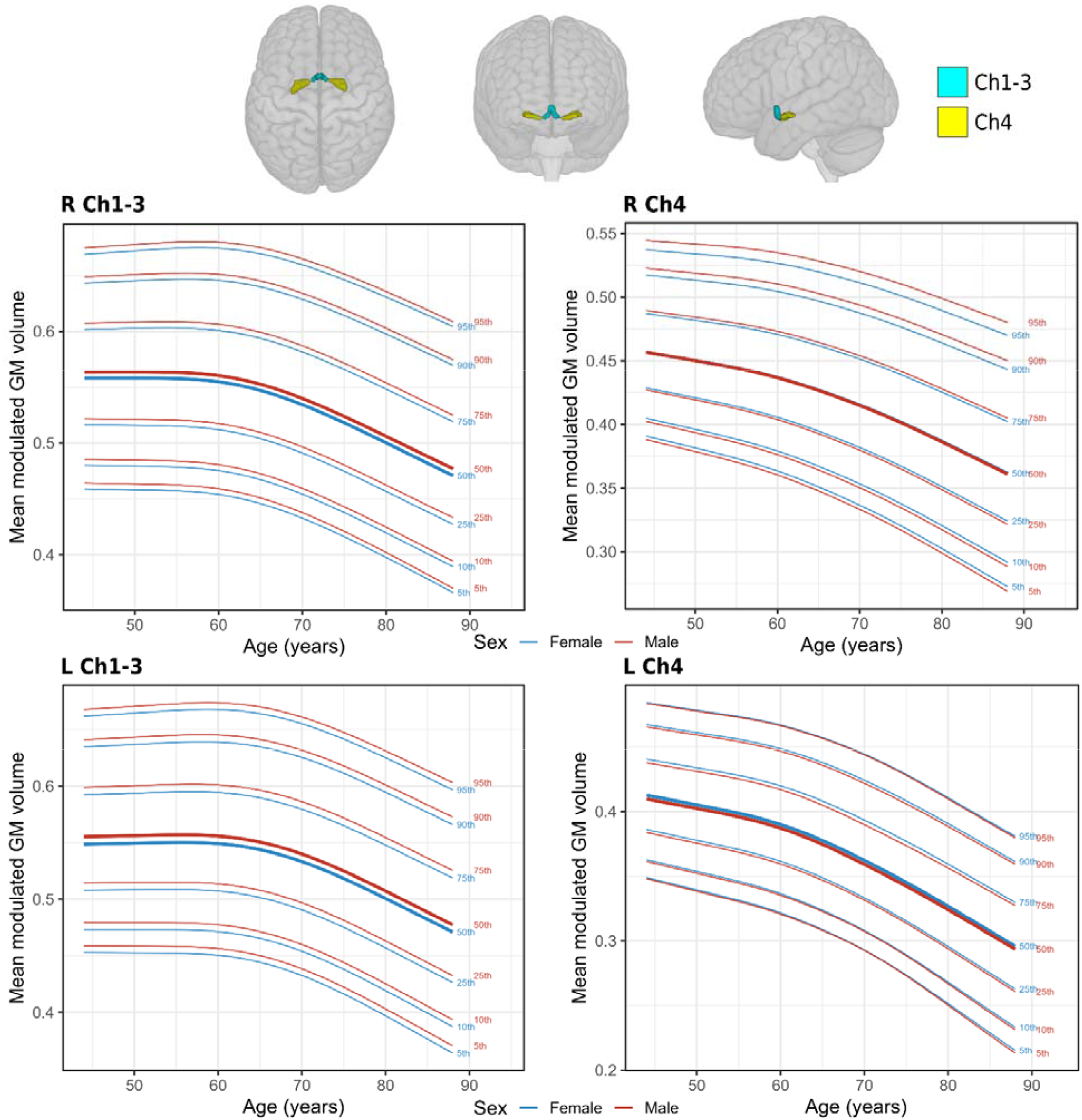
Basal forebrain compartments GM volume normative trajectories from mid to late life. Population centile curves for grey-matter volume in the septal/diagonal-band compartment (Ch1–3, left) and nucleus basalis of Meynert (Ch4, right), shown separately for the right (top) and left (bottom) hemispheres, from generalised additive models for location, scale and shape (GAMLSS) fitted on a healthy reference sample of UK Biobank participants aged 45–87 years. Curves show the 5th, 10th, 25th, 50th, 75th, 90th and 95th centiles as a function of age, modelled separately by sex (blue, female; red, male). Ch4 volume declines with age from the start of the sampled range in both hemispheres, whereas Ch1–3 volume is relatively stable until the seventh decade before declining. L = left; R = right

### Normative modelling of basal forebrain structural integrity

Normative modelling charts the expected distribution of a brain measure as a function of age and sex, and re-expresses each individual’s value as a deviation from that expectation^21,23^. To assess the macrostructural integrity of the BF subnuclei across the adult lifespan and test whether the Ch4 and Ch1–3 subsystems follow distinct normative trajectories through ageing, we used generalised additive models for location, scale and shape (GAMLSS)^22,40^ implemented in the *gamlss* package in R^22,40^, enabling to capture non-linear age trajectories, heteroscedasticity, and potential non-normality. GAMLSS allows flexible modelling of multiple distribution parameters beyond the mean, permitting age-related shifts in variance, skewness, and kurtosis. Trajectories were estimated in a healthy reference sample (n = 43,586-43,620, depending on the specific deconfounded BF metric) comprising participants without major neurological, psychiatric, or systemic diagnoses recorded either before imaging or during the available UK Biobank follow-up. The full list of exclusion criteria is provided in **Supplementary Table 1**.

We modelled the grey matter volumes of the Ch1-3 and Ch4, estimated separately for the left and right hemispheres, and the two hemispheric derived z-scores were then averaged to give the single bilateral measure for each BF sub-component which was then used in downstream analyses. Non-linear effects of age across all four parameters were modelled using Penalized B-splines (pb), where smooth function degrees of freedom were automatically selected via generalized cross-validation (GCV) to balance model flexibility and guard against overfitting. Main effects for sex were included across all four parameters to account for sex-specific baseline differences in location (μ) and scale (σ) skewness (ν) and kurtosis (τ). To control for site-specific variance across the multi-centre data without overfitting, random intercepts for site were included for location and scale. For each BF metric the distributional family was selected from a candidate set of continuous distributions, which were empirically tested to identify the optimal parametric form for each BF metric. Model performance and distribution family selection were assessed based on two complementary criteria: 1. Model stability – determined by successful convergence within the default log-likelihood threshold of 0.001 between iterations; 2. The Bayesian information criterion (BIC) (**Supplementary Table 2)**. Individual deviations from the normative trend (Z-scores) were derived using quantile randomized residuals, mapping fitted distribution quantiles onto a standard Gaussian scale to reflect the number of standard deviations from the normative median. To ensure model robustness, normative parameters (μ, σ, ν, τ) were estimated using 20-fold cross-validation^41,42^. In each iteration, models were trained on 19 folds and applied to the remaining “unseen” test fold to estimate centiles and corresponding deviation scores. The deviation scores for participants outside the healthy reference sample were predicted from the normative model fitted in the healthy reference sample. Scores are oriented so that higher values indicate greater-than-expected GM volume for age and sex while negative values indicate lower-than-expected.

### Basal forebrain grey matter volume change over time

To examine whether the age-related patterns observed cross-sectionally are also evident longitudinally, we assessed change in BF compartment GM volume over time using a subset of participants from the healthy reference sample with follow-up imaging data (n = 2,712). For this analysis we used the deconfounded (i.e., cleaned) raw grey matter volumes (i.e., the same that were entered to the normative models) and not the normative deviation scores. This analysis followed previous works^43,44^, and was conducted using generalized additive mixed models (GAMMs) implemented with the *gamm4* package in R. In order to separate cross-sectional and longitudinal effects of age, we decomposed age into 1) ***mean-age***, representing the cross-sectional effect of age by calculating the average age across time points for each participant, and 2) ***time***, representing the longitudinal effect of age by calculating the deviation of each participant’s age at a given time point from their mean-age^43^. Mean-age was modelled as a smooth spline with a maximum of 20 basis functions. Time was modelled as a linear term due to the relatively short within-person follow-up period. Sex was modelled as a binary factor. The models also included random intercepts for UK Biobank assessment centre and participant ID. To examine whether the rate of Ch1-3 or Ch4 grey matter volume changed over time as a function of age, a linear time x mean-age interaction term was evaluated in the model. Then, to directly examine whether time x age trends were compartment-dependent, we fitted a model with a three-way time x mean-age x BF compartment interaction, with all lower-order interactions included.

### Basal forebrain structural vulnerability in neuropsychiatric disorders

We next tested whether normative deviations of the basal forebrain subnuclei, the Ch4 and the Ch1–3, show vulnerability to major neurodegenerative and psychiatric disease aetiologies using three complementary lines of evidence: (i) case–control comparison of manifest disease, (ii) case–control comparison of pre-diagnostic disease and prospective association with incident diagnosis, and (iii) association with disease-related polygenic risk scores. The outcomes throughout these lines of analysis were the normative deviation (i.e., the age-gap z-scores) of the Ch4 and Ch1–3 GM volumes.

#### Case-control structural vulnerability in manifest neuropsychiatric disorders

Manifest disease groups were defined based on information obtained from UK Biobank linked primary care records, hospital inpatient data, death records, and self-reported medical conditions, mapped to ICD-10 diagnostic codes, following an approach recommended by UK Biobank^45^. Therefore, the disease groups were defined *a priori* from recorded diagnoses present on or before the imaging visit for: all-cause dementia, Parkinson’s disease, other parkinsonism, multiple sclerosis and demyelinating disease, schizophrenia, bipolar disorder, recurrent depression and stroke and cerebrovascular disease (see **Supplementary Table 1** for diagnostic codes). For this analysis, only cases with mutually exclusive diagnoses were included for clearer interpretation (i.e., without comorbidities within this group of diagnoses). Each disease group was then compared against the healthy reference subsample. For each disease × outcome, we fitted a generalised additive model, using the *mgcv* package in R, of the outcome on case status, adjusting for age (penalised thin-plate spline), sex, Townsend deprivation index, educational attainment, smoking status (dichotomised as ever vs never), alcohol-intake frequency, and body mass index (BMI), with imaging site as a random intercept (see **Supplementary Table 3** for all covariates’ UK Biobank data fields). Age and Townsend deprivation scores were modelled as penalised thin-plate splines smooth terms. The adjusted GAM β (unstandardized, in normative-z units) was reported as the primary effect size. Confidence intervals and *p*-values were obtained analytically from the fitted model (Wald intervals, β ± 1.96 × SE). Statistical significance was determined by the covariate-adjusted model and controlled for multiple comparisons across the full disease × outcome table using the Benjamini–Hochberg false discovery rate (FDR)^46^. Furthermore, to test the anatomical dissociation directly — rather than inferring it from the separate significance of two models — we fitted linear mixed models using the *lme4* package in R in which each participant contributed both the Ch4 and Ch1–3 metrics and the model was constructed as outcome ∼ case × subnucleus + age + sex + covariates + (1 | site) + (1 | participant), assessing UKB site and participants as random intercepts. The case × subnucleus interaction was the quantity of interest, estimating the within-disease difference in the case effect between the BF compartments. Models were fitted on participants with complete data for both compartments, so the compared structures were estimated in the same individuals. Interaction terms were corrected for multiple comparisons across diseases using FDR.

#### Case-control structural vulnerability in pre-diagnostic neurodegeneration

First, to determine whether BF deviation is already detectable before clinical diagnosis of the major neurodegenerative conditions, namely, all-cause dementia and PD, we compared participants who were free of all the major neuropsychiatric conditions at imaging and subsequently received a first diagnosis of dementia or PD, against the healthy reference sample, similarly to the case-control procedure. Diagnoses occurring within 12 months of imaging were excluded, removing individuals plausibly already symptomatic or under investigation at the time of scanning. We first ran a primary analysis in which pre-diagnostic cases were defined as disease-free at baseline with non of the examined major neuropsychiatric conditions diagnosed at imaging. Then we ran a sensitivity analysis with a “Target-first” definition, with no major neuropsychiatric conditions diagnosed at imaging and the target diagnosis (dementia or PD) must be the first to be diagnosed among all other listed disorders. The only exception was for PD, where other parkinsonism diagnosis within the year after the imaging visit were permitted, given the known potential for diagnostic reclassification in parkinsonian disorders^47^. The same case × subnucleus interaction models were fitted to test the anatomical dissociation directly in the pre-diagnostic groups. The same was performed for

#### Prospective risk of incident neurodegenerative disease (time-to-diagnosis analysis)

Next, we tested whether baseline cholinergic integrity predicted incident all-cause dementia and PD using Cox proportional-hazards regression. Participants with any of the studied neuropsychiatric conditions manifest at imaging visit were excluded from this analysis. Follow-up ran from the imaging visit to the earliest of first diagnosis, death, or the UKB censoring date of 31 March 2023. Participants were therefore right-censored at death or administrative end. To separate a basal forebrain–specific association from confounding by other general and more specific risk factors, we ran five sets of pre-specified models. 1) a minimally adjusted model (age, sex) was examined; 2) a general-covariates model adjusted for age, sex, Townsend deprivation, education, smoking status, alcohol-intake frequency, BMI; 3) a general-covariates model adding a relevant PRS (i.e., AD or PD); 4) a general-covariates model adding generalised disease-specific brain markers: hippocampal and cortical grey-matter normative deviations for dementia, and substantia nigra magnetic susceptibility deviation for PD^48^, each expressed on the same normative-deviation scale after initial deconfounding of the raw metrics; 5) a fully-adjusted model with all variables. The individual deviation scores for the disease-specific brain metrics were derived through the same normative modelling procedure as the BF deviation age-gap scores (see **Supplementary Table 4** and **Supplementary Figure 1**). In addition, in all models, Ch4 and Ch1–3 were each entered either alone or jointly together, to distinguish each compartment’s independent contribution from their shared signal. Covariate completeness was required for each of the models, so the analytic sample and cases numbers vary across sets (**Supplementary Table 5**). Hazard ratios are expressed per unit of normative deviation, oriented so that higher values denote better integrity. Hazard ratios for the four metrics were FDR-corrected within each disease. The proportional-hazards assumption was assessed for every term in every model using scaled Schoenfeld residuals, both per term and globally. The assumption was supported for all terms except Ch4 in the dementia models, where it was violated in most models. A violation indicates that the estimated hazard ratio is a time-averaged summary of an effect that varies over follow-up. Therefore, the time-averaged hazard ratio is reported as the primary estimate for comparability with the other outcome and compartments, and the time-dependence is characterised directly in a pre-specified sensitivity analysis (below). To characterise the proportional-hazards violation of Ch4 in the dementia models, we fitted continuous time-varying-coefficient Cox models. The coefficient of each BF term was allowed to vary with time as β(t) = β + β ·log(t), implemented through the time-transform (tt) mechanism of the *survival* package in R. This parameterisation yields a baseline log-hazard ratio (β) and a slope on log-time (β) whose sign indicates whether the association strengthens (β < 0) or attenuates (β > 0) over follow-up. Models were fitted across the same covariate ladder, for Ch4 entered alone and jointly with Ch1–3.

#### Polygenic risk for neuropsychiatric disorders and BF structural integrity

To estimate the association between genetic liability and the structural vulnerability of the BF we used polygenic risk scores (PRS) from UK Biobank’s standard PRS release for Alzheimer’s disease, Parkinson’s disease, multiple sclerosis, schizophrenia, bipolar disorder and ischaemic stroke^49^. To obtain a genotype–phenotype association unconfounded by prevalent or emerging disease, we restricted this analysis to participants with no recorded history — past or subsequent — of any of the studied neurodegenerative, cerebrovascular or psychiatric conditions. For each polygenic risk score × outcome (BF compartment), we fitted a generalised additive model of the outcome, adjusting for age as a smooth term, sex, and imaging site (random effect). To further adjust for population structure, we also added the first ten genetic principal components to all models. We did not adjust for additional covariates, i.e., lifestyle or socioeconomic, as these are not expected to have an effect on participants genotype and to confound the associations^50^. The *p*-values of all PRS predictors across the models were corrected with FDR.

### Sensitivity analyses

We performed several sensitivity analyses to complement the primary findings. First, while a 50% threshold was shown to be in accordance with realistic histology-based volume estimation of the Ch4/NbM^37^, such was not examined in the Ch1-3 compartment. We therefore re-ran the normative modelling and GM volume change over time analyses using 30%-thresholded masks of the right and left Ch1-3, to examine whether the patterns converge and are therefore generally independent of the atlas threshold selection. Second, because the primary case–control comparison estimated the case effect by covariate adjustment across the full reference sample, we ran a matched sensitivity analysis to confirm the effect was not driven by residual covariate imbalance or by the functional form of the adjustment. For each condition, cases were matched to disease-free reference participants by 1:5 nearest-neighbour propensity-score matching without replacement, with a calliper of 0.2 propensity-score standard deviations. The propensity score was estimated from age, sex, and the covariates used in the primary analysis models. Covariates that did not vary within a given condition’s sample were dropped from that condition’s propensity model. The case-effect model was then refitted within each matched set using the same specification as the primary analysis, and FDR-correction was applied across conditions as before. The same was also performed for the pre-diagnostic case-control analysis. Lastly, the primary pre-diagnostic case-control analysis required the target diagnosis to occur at least 12 months after imaging, to exclude individuals plausibly already symptomatic or under investigation at the scan. To test whether the pre-diagnostic deficit was robust to this choice and not an artefact of including cases diagnosed shortly after imaging, we re-ran the primary case–control fingerprint at progressively longer definition cut-offs of 12, 24, 36, and 48 months, and characterised how the effect behaves as the cases nearest to diagnosis are progressively removed.

### Statistical analysis

Data processing and statistical analyses were conducted in R (version 4.4) and RStudio (version 2024.04.2).

## Results

### Normative modelling of basal forebrain cholinergic subnuclei across mid-to-late adulthood

We first constructed normative reference models for the two principal compartments of the basal forebrain cholinergic system — Ch1–3 (medial septum and vertical/horizontal limbs of the diagonal band) and Ch4 (NbM) — in participants across mid-to-late life between the ages of 45 and 87 years. Quality-control diagnostics of the models are presented in **Supplementary Figure 2**. The fitted median trajectories revealed distinct age-related patterns across the two basal forebrain sub-systems, suggestive of different age-related profiles across this age range (**Figure 1**). Bilateral Ch1–3 volume remained relatively stable between approximately 45 and the late 50s, followed by a nonlinear decline that became progressively steeper from approximately 60–65 years onward. A similar pattern was observed when a 30% threshold was used to define the Ch1-3 masks (**Supplementary Figure 3**). In contrast, bilateral Ch4 volume declined throughout the observed age range, with increasingly pronounced reductions at older ages. The separation between the lower and upper centile curves generally increased with age in both regions, indicating increasing estimated interindividual variability in basal forebrain volume at older ages. The increasing separation of the normative centiles with advancing age was supported by positive age effects on the scale parameter in all four models. For Ch1–3, the age effects on scale were β = 0.00249–0.00294 (both *p* < 0.001), while corresponding effects for Ch4 were β = 0.00441– 0.00680 (both *p* < 0.001), indicating increasing conditional variability in grey-matter volume with age across both basal forebrain compartments. Sex differences in bilateral Ch1–3 were primarily expressed as a small shift in the central location of the volume distribution, with males showing higher conditional volumes than females in both hemispheres (β = 0.00544– 0.00639, both *p* < 0.001). In contrast, there was no evidence for sex-related differences in Ch1–3 dispersion (β = −0.00049 to 0.00145, *p* = 0.837–0.944), consistent with the broadly parallel male and female centile trajectories observed in **Figure 1**. For Ch4, sex differences in central volume were small, with sex effect on the location parameter ranging from β = −0.00210 to 0.00085 per hemisphere. Instead, the clearest bilateral sex-related difference in Ch4 concerned distributional skewness: males showed higher skewness parameters in both hemispheres (β = 0.399–0.485, both *p* < 0.001). Thus, although the male and female central Ch4 trajectories were generally closely aligned, sex differences were more apparent in the shape of the conditional volume distribution than in its central tendency.

### Change in the basal forebrain compartments grey matter volume over time

We then tested the patterns of longitudinal change in GM volume of the BF compartments over time using individuals from the healthy reference sub-sample with follow-up imaging data (n = 2,712, 2.6±1.10 years interval, range 1.00-7.17 years). While both Ch4 and Ch1-3 showed a decline over time (Ch4: β = −0.0005 [−0.0009, −0.00012], *p* = 0.010; Ch1-3: β = −0.0006 [−0.001, −0.00005], *p* = 0.033), only the Ch1-3 demonstrated a time x age interaction (Ch1-3: β = −0.0002 [−0.0003, −0.0001], *p* < 0.001; Ch4: *p* = 0.864) (**Figure 2**).

**Figure 2.**
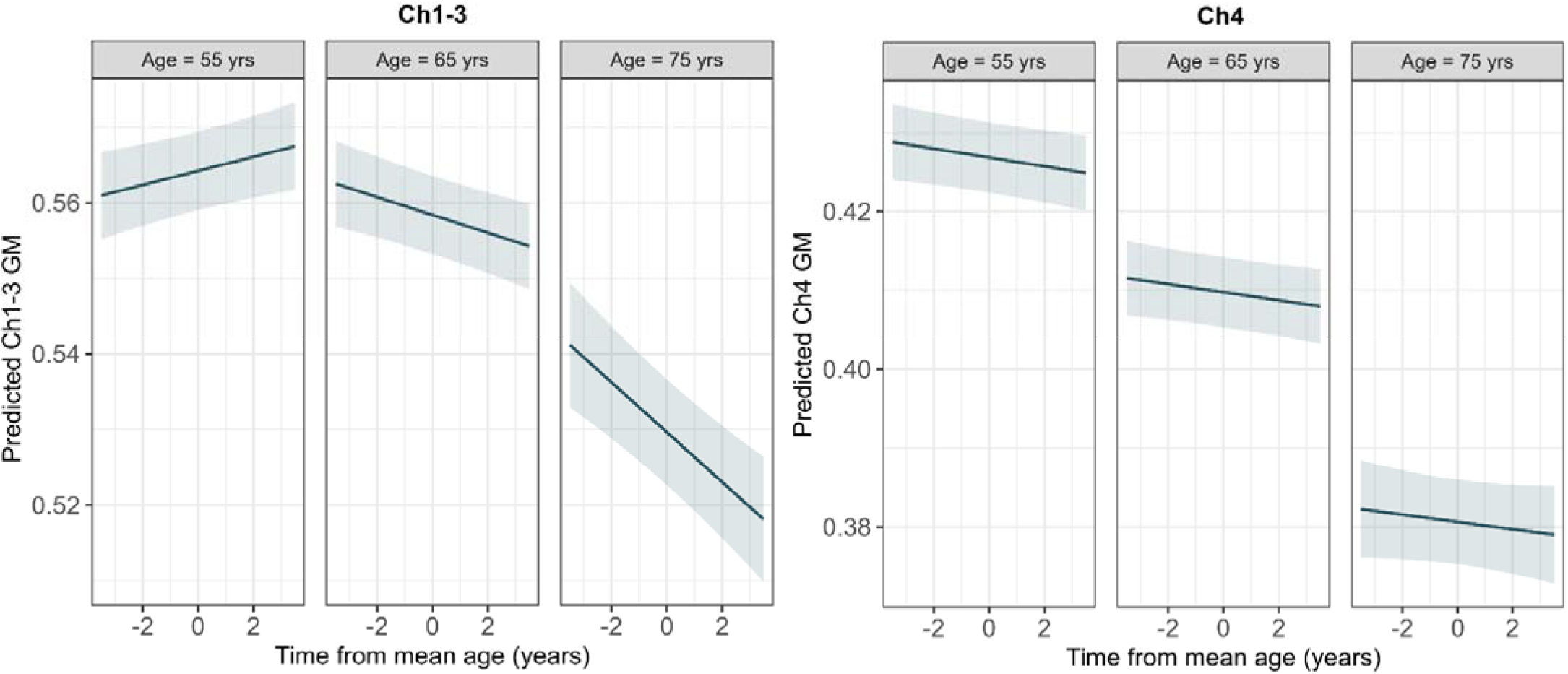
Distinct basal forebrain compartments GM volume change over time. Distinct age-related changes over time in grey matter volume were observed between Ch1-3 (left) and the Ch4 (right) using generalized additive mixed models (GAMM) performed on a subset of the healthy reference sample with two time points of imaging data (n=2,712). The model-based predicted grey matter volume changes are presented for three different ages (55, 65, 75 years) showing that Ch1-3 decline from the seventh decade while a persistent decline from mid-life is already observed for Ch4 grey matter volume.

This differential pattern was further supported when we directly tested a time x age x compartment interaction in GM volume change over time (β = 0.0002 [0.0001, 0.0003], *p* = 0.004), adhering to the differential cross-sectional trajectories of the two compartments. A similar pattern for the Ch1-3 was observed with a 30% threshold masks (**Supplementary Figure 4**).

### Deviation in manifest neurological disease is generally concentrated in Ch4

We next asked whether individual deviations from the normative expected trajectories distinguish participants with diagnosed neuropsychiatric disorders at the time of scanning. Negative deviations (i.e., less-than-expected GM volume) were found in all five neurological conditions examined compared to the healthy reference sub-sample (n = 42,032/42,067 for Ch1-3 and Ch4 respectively) (**Figure 3A**). Furthermore, while all-cause dementia, MS/demyelinating disease, and cerebrovascular disease showed significant decline in both BF compartments, PD and other parkinsonian disorders were only characterized with reduced Ch4: *all-cause dementia* (n = 43): Ch4 β = −0.605 [−0.861, −0.350], *p*FDR < 0.001, Ch1–3 β = −0.354 [−0.627, −0.081], *p*FDR = 0.022; *multiple sclerosis / demyelination* (n = 291): Ch4 β = −0.558 [−0.656, −0.459], *p*FDR < 0.001, Ch1–3 β = −0.175 [−0.280, −0.069], *p*FDR =0.003; *stroke / cerebrovascular disease* (n = 1,394): Ch4 β = −0.203 [−0.249, −0.156], *p*FDR < 0.001, Ch1–3 β = −0.187 [−0.237, −0.138], *p*FDR < 0.001; *Parkinson’s disease* (n = 162): Ch4 β = −0.319 [−0.451, −0.187], *p*FDR = 0.001, Ch1–3 β = +0.088 [−0.052, 0.229], *p*FDR = 0.32; *other parkinsonism*: Ch4 β = −0.098 [−0.169, −0.028], *p*FDR = 0.014; Ch1–3 (n = 572; β = −0.051 [−0.127, 0.024], *p*FDR = 0.291. No psychiatric condition showed statistical evidence of altered BF structural integrity in either structure (**Figure 3A**): *recurrent depression* (n = 329; Ch4 β = −0.016 [−0.110, 0.076], *p*FDR = 0.789; Ch1-3 β = −0.052 [−0.151, 0.047], *p*FDR = 0.373), *bipolar disorder* (n = 175; Ch4 β = −0.101 [−0.227, 0.026], *p*FDR = 0.213; Ch1-3 β = −0.078 [−0.214, 0.058], *p*FDR = 0.346) and *schizophrenia* (n = 51; Ch4 β = −0.041 [−0.274, 0.194], *p*FDR = 0.787; Ch1-3 β = 0.001 [−0.250, 0.252], *p*FDR = 0.993). To further test the specificity of the affected BF compartments within participants in each disease rather than only across disease groups, a case × compartment interaction was estimated within-subjects with mixed models (**Figure 3B**). The interaction was statistically significant for two conditions, namely PD and MS/demyelinating disease, both showing statistical evidence for Ch4 GM in integrity being more reduced (*PD*: case × compartment β = 0.399 [0.236, 0.562], *p*FDR < 0.001; *MS/demyelinating disease:* case × compartment β = 0.380 [0.257, 0.502], *p*FDR < 0.001). All-cause dementia showed the same direction at similar magnitude but was not significant after multiple comparisons correction (case × compartment β = 0.237 [−0.080, 0.553], *p*FDR = 0.38), consistent with the modest 43-case sample size rather than with an absent effect. No neuropsychiatric condition in this dataset showed a larger statistically significant reduction in Ch1–3 than in Ch4. The case-control between-group differences were similar under matching (**Supplementary Figure 5A**), indicating the primary associations were not artefacts of covariate imbalance.

**Figure 3.**
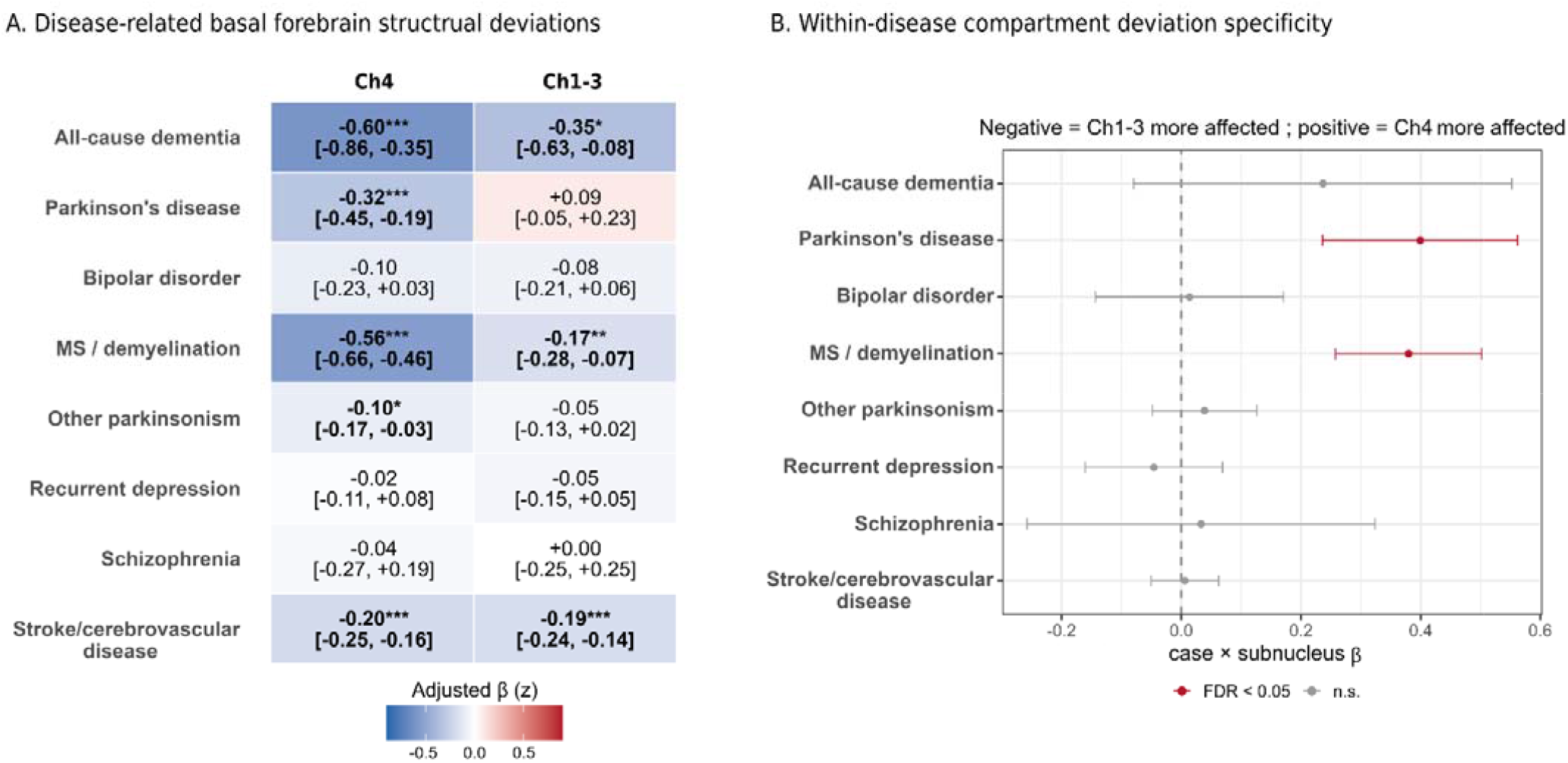
Structural vulnerability of the basal forebrain compartments in neurological and psychiatric disorders. (A) Adjusted case–control differences in normative deviation (β, in age- and sex-conditioned z-units; 95% CI) for the nucleus basalis (Ch4) and the septal/diagonal-band compartment (Ch1–3) across eight conditions, from covariate-adjusted generalised additive models (Methods). Negative values (blue) indicate lower-than-expected grey-matter volume for age and sex; colour encodes effect magnitude and direction, and asterisks with bold type denote associations surviving FDR-correction (* *p*FDR *< 0.05, ** p*FDR *< 0.01,* *** *p*FDR < 0.001). Ch4 showed negative deviation across all five neurological conditions, numerically larger than the corresponding Ch1–3 deviation, whereas no psychiatric disorder (bipolar disorder, recurrent depression, schizophrenia) showed statistical evidence of deviation in either compartment. (B) Within-disease compartment specificity, tested as a case × subnucleus interaction (β with 95% CI); positive values indicate a larger deficit in Ch4, negative a larger deficit in Ch1–3. The Ch4-greater-than-Ch1–3 difference was significant for PD and MS/demyelination (*p*FDR < 0.05). All-cause dementia showed the same direction, but the difference did not survive correction; the modest number of dementia cases (n = 43) likely limited power for this within-disease contrast. MS = multiple sclerosis; n.s. = non-significant (FDR-corrected); PD = Parkinson’s disease

### Reduced Ch4 structural integrity is present years before diagnosis

We then restricted analysis to participants who were free of all the major neuropsychiatric disorders at the time of imaging and received a first diagnosis of either dementia (n=126) or PD (n=70) at least 12 months afterwards, compared against the same healthy reference sample. Median time from scan to diagnosis was 4.6 years (IQR 3.6-6.7; range 1.1–9.2) for dementia and 4.1 years (IQR 3.0–5.6; range 1.0–8.6) for PD. Below normative Ch4 deviation was found in both groups: dementia β = −0.560 [−0.709, −0.412], *p*FDR < 0.001; PD β = −0.358 [−0.558, −0.158], *p*FDR < 0.001. Ch1–3 showed a more modest deviation in the pre-diagnostic dementia cases (β = −0.217 [−0.377, −0.058], *p*FDR = 0.010) with no statistical evidence for deviation from healthy individuals before PD diagnosis (β = 0.128 [−0.086, 0.342], *p*FDR = 0.240) (**Figure 4A**). Furthermore, a significant interaction was found in both groups for greater deviation from normal trajectories in the Ch4 compared to Ch1-3 (dementia: β = −0.342, *p*FDR < 0.001; PD: β = −0.485, *p*FDR < 0.001).

**Figure 4.**
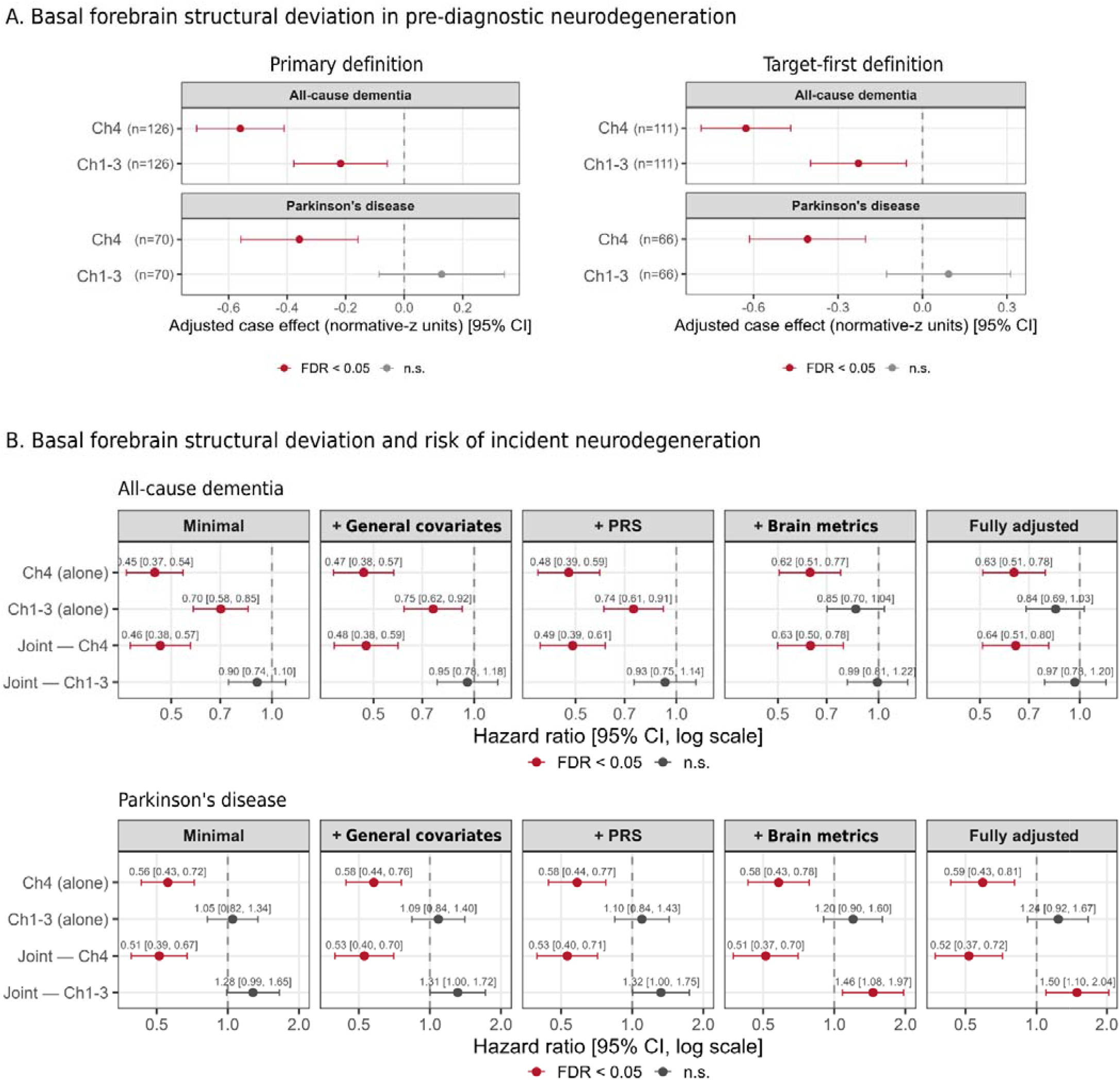
Basal forebrain structural deviation in pre-diagnostic stages of neurodegeneration. (A) Pre-diagnostic deviation. Adjusted case–control differences in deviation (normative-z units, 95% CI) between participants who received a first diagnosis of all-cause dementia or Parkinson’s disease at least 12 months after imaging and a disease-free reference sample, estimated from covariate-adjusted generalised additive models (age, sex, deprivation, education, smoking, alcohol, body-mass index). The primary definition (left) requires only that the target diagnosis post-dates the imaging by at least 12 months; the target-first definition (right) additionally requires the target to be the first diagnosis recorded after imaging among all other neuropsychiatric conditions (Methods). Case numbers are shown per row; more negative values indicate lower-than-expected volume in future cases. (B) Prospective prediction. Cause-specific Cox proportional-hazards ratios for incident all-cause dementia (top) and Parkinson’s disease (bottom), per +1 normative-z; a hazard ratio below 1 indicates that greater-than-expected volume is associated with lower incidence. Each compartment was entered alone (Ch4 alone; Ch1–3 alone) and jointly (Joint — Ch4; Joint — Ch1–3), across five nested covariate sets: minimal (age, sex); + general covariates (deprivation, education, smoking, alcohol, body-mass index); + polygenic risk; + reference brain metrics (hippocampal and cortical deviation for dementia; substantia nigra deviation for PD); and fully adjusted (all covariates together). Hazard ratios and 95% confidence intervals are annotated per estimate. CI = confidence interval; n.s. = non-significant (FDR corrected); PD = Parkinson’s disease; PRS = polygenic risk score

When restricting the future case groups to a target-first definition (dementia: n=111, PD: n = 66), similar results were observed with Ch4 deviation demonstrated in both groups: dementia β = −0.627 [−0.786, −0.468], *p*FDR < 0.001; PD β = −0.408 [−0.614, −0.202], *p*FDR < 0.001, and Ch1–3 deviation observed in the pre-diagnostic dementia cases (β = −0.227 [−0.398, −0.057], *p*FDR = 0.012) but no in the pre-diagnostic PD group (β = 0.093 [−0.128, 0.313], *p*FDR = 0.410) (**Figure 4A**). A significant interaction was also found under the target-first definition for greater deviation of Ch4 compared to Ch1-3 in both groups (dementia: β = −0.400, *p*FDR < 0.001; PD: β = −0.501, *p*FDR < 0.001). The case-control between-group differences were essentially unchanged under matching (**Supplementary Figure 5B**). Furthermore, the pre-diagnostic Ch4 deficit was generally stable across the cut-off definitions (β = −0.56, −0.56, −0.51, and −0.43 across the 12, 24, 36, and 48 months, respectively, *p*FDR < 0.001, **Supplementary Figure 6**), despite the case count falling from 126 to 79. For PD the Ch4 deficit was likewise stable across the 12-, 24-, and 36-month cut-offs (β = −0.36, −0.33, −0.35; *p*FDR ≤ 0.006) and lost statistical significance only at the 48-month cut-off, where the estimate remained negative but the case count had fallen to 37 (β = −0.19, *p*FDR = 0.34), consistent with reduced power rather than an attenuating effect (**Supplementary Figure 6**). In terms of the Ch1–3 compartment, its modest dementia association at the 12-month definition (β = −0.22, *p*FDR = 0.010) attenuated to the null at 48 months (β = −0.11, *p*FDR = 0.40 at), and its PD association was null at every cut-off definition.

### Baseline Ch4 deviation is linked to incident dementia and Parkinson’s disease

Among participants free of the relevant condition at scanning (n = 53,479; median 4.2 years of follow-up time, 129 incident dementia and 76 incident PD diagnoses), greater baseline negative deviation of Ch4 was associated with higher incidence of both diseases, and the association survived every level of adjustment **(Figure 4B).** For dementia, the Ch4 hazard ratio per +1 normative-z was 0.45 [0.37, 0.54] in the minimal model, 0.47 [0.38, 0.57] with further adjustment for deprivation, education, smoking, alcohol and body-mass index, 0.48 [0.39, 0.59] with the addition of PRS, and 0.62 [0.51, 0.77] with hippocampal and cortical deviation additionally included (all *p*FDR < 0.001). For PD the corresponding Ch4 hazard ratios were 0.56 [0.43, 0.72], 0.58 [0.44, 0.76], 0.58 [0.44, 0.77], and 0.58 [0.43, 0.78] (all *p*FDR p < 0.001). Adjustment for hippocampal and cortical deviation attenuated the dementia association—consistent with a component of shared, generalised atrophy—but did not abolish it; the PD association was essentially unchanged by adjustment for nigral susceptibility deviation.

When entered alone, Ch1–3 deviation also predicted incident dementia (minimal HR 0.70 [0.58, 0.85], *p*FDR < 0.001), but this association did not survive joint modelling with Ch4: in the same model, the Ch1–3 hazard ratio moved to 0.90 [0.74, 1.10] (minimal), 0.95 [0.78, 1.18] (covariate + PRS adjusted), and 0.99 [0.81, 1.22] (with reference brain metrics), indistinguishable from the null throughout (all *p*FDR > 0.30), while Ch4 retained its association in the same joint models (e.g. minimal 0.46 [0.38, 0.57], *p*FDR < 0.001). For PD, Ch1–3 showed no association in any single-metric model (minimal HR 1.05 [0.82, 1.34], FDR p = 0.71). Note that in the most heavily adjusted joint models in PD, the Ch1–3 hazard ratio rose above unity and reached nominal significance (e.g., fully adjusted + reference brain 1.46 [1.08, 1.97]). However, this may likely be a result of statistical suppression by a correlated Ch4 term at a low events-per-variable ratio rather than a protective effect of lower Ch1–3 volume.

In dementia, Ch4 violated the proportional-hazards assumption, whereas no other term violated it in any model. To characterise this, we ran a sensitivity analysis with a continuous time-varying-coefficient model. In the joint models, in which Ch4 and Ch1–3 were entered together, the Ch4 coefficient carried a positive slope on log-time — indicating that its protective association with incident dementia was strongest shortly after imaging and attenuated toward the null over follow-up (**Supplementary Figure 7**). In the minimal joint model the Ch4 time-varying hazard ratio was 1.63 [1.15, 2.33] per unit log-time (β = 0.49; *p*FDR = 0.034), against a Ch1–3 slope indistinguishable from the null (1.16 [0.84, 1.59], p = 0.38). This pattern persisted under adjustment for general covariates (Ch4 1.55 [1.05, 2.31], *p*FDR = 0.049) and PRS (1.59 [1.07, 2.35], *p*FDR = 0.043) and was directionally unchanged but no longer statistically significant once hippocampal and cortical deviation were included (1.36 [0.93, 2.00], *p*FDR = 0.11). Throughout, the Ch1–3 time-varying coefficient remained small and non-significant (all *p* ≥ 0.38) and its baseline coefficient did not differ from the null (all *p* ≥ 0.22), indicating that neither an independent prospective association nor its time-dependence was present in the Ch1-3 compartment.

### Neuropsychiatric genetic liability association with basal forebrain structural integrity

Standardised polygenic risk scores showed weak associations with basal forebrain integrity in a sample free from any history of the major neuropsychiatric disorders (n = 72,019 for Ch4; n = 71,957 for Ch1–3) (**Figure 5A**). Monotonic and generally linear associations were found between lower structural integrity of the Ch4 and higher PRS for AD (edf = 1.57, *p*FDR < 0.001), bipolar disorder (edf = 1.03, *p*FDR < 0.001), MS/demyelinating-disease (edf = 1.68, *p*FDR < 0.001), and schizophrenia (edf = 1.00, *p*FDR < 0.001), but no relationship was observed with the PRS of PD (edf = 1.00, *p*FDR = 0.570) or ischemic stroke (edf = 1.00, *p*FDR = 0.993). Lower structural integrity of the Ch1-3 was associated with higher PRS for AD (edf = 1.01, *p*FDR < 0.001), ischemic stroke (edf = 1.00, *p*FDR < 0.001), schizophrenia (edf = 1.00, *p*FDR < 0.001), bipolar disorder (edf = 2.24, *p*FDR < 0.001), and MS/demyelinating-disease (edf = 2.34, *p*FDR = 0.040), but not for PD (edf = 1.00, *p*FDR = 0.145). Nevertheless, no model exceeded a partial R² of 0.0005, suggesting that very small amounts of variance in BF integrity explained by each of the PRS metrics. Only ischemic stroke PRS showed a compartment-specific interaction that survived FDR correction, with greater link to Ch1-3 structural integrity, although the effect size itself was very small (**Figure 5B**).

**Figure 5.**
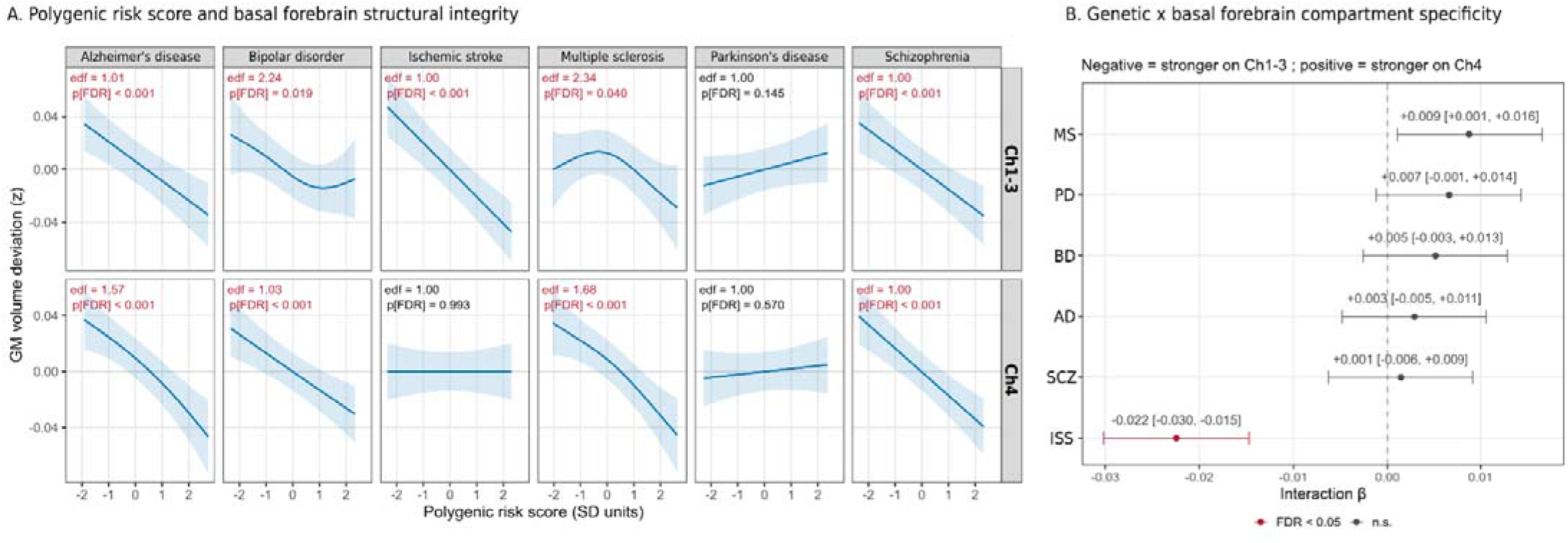
Polygenic liability shows small, largely non-specific associations with basal forebrain compartments structural deviations. (A) Penalised-spline associations between each of the six available neuropsychiatric polygenic risk scores and structural deviation in Ch1–3 (top) and Ch4 (bottom), derived from generalised additive models; shaded bands are 95% confidence intervals. Effective degrees of freedom (edf) are shown per panel (edf ≈ 1 indicates a mainly linear relationship), and FDR-corrected values are presented in red. Although several associations are statistically significant at this sample size, all are of small magnitude as can be appreciated by the y-axis range spanning a small fraction of the deviation-score distribution. (B) Compartment specificity of each polygenic association, tested as a PRS × compartment interaction (β with 95% CI); positive values indicate a stronger association with Ch4, negative with Ch1–3. Interaction models were fitted with age, sex, and ten genetic principal components as covariates (see Methods). FDR-corrected values are presented in red. AD = Alzheimer’s disease; BD = bipolar disorder; edf = effective degrees of freedom; GM = grey matter; ISS = ischemic stroke; MS = multiple sclerosis; PD = Parkinson’s disease; SCZ = schizophrenia; SD = standard deviations

## Discussion

In this study we aimed to examine whether the two principal compartments of the human BF—the NbM/Ch4 and the septal-diagonal/Ch1–3—differ in how they age across mid-to-late adulthood and in their structural vulnerability to neurological and psychiatric disease. Using population-scale normative models we found that the two compartments follow distinguishable profiles across this age range, evident from both the cross-sectional and within-subject changes over time, suggesting against a uniformly ageing system. While Ch4 was found to demonstrate a decline across the age range, Ch1-3 volume was relatively stable until the seventh decade of life. These dissociable profiles are supported by a previous study in healthy adults aged 20–94 years, showing the most pronounced linear age-related decline in the Ch4^19^. It has been proposed that the BF vulnerability cannot be solely explained by downstream pathology consequences but may also reflect intrinsic features relating to their evolutionary and developmental properties, and functional demands across the lifespan^18^. For instance, the NbM shows an evolutionary expansion across species which parallels that of the neocortex, likely reflecting the increasing demand of cholinergic signalling needed to support the progressively more complex cognitive functions^51,52^. Further, these requirements, along with the physical properties of these neurons and their unmyelinated axonal projections, are also associated with increased bioenergetic demands and metabolic activity, making them more vulnerable to high levels of cellular stress^53–55^.

Negative Ch4 deviations were more pronounced in neurological conditions compared to the Ch1-3, and statistically more extreme in PD and MS/demyelinating disease. Dementia showed the same pattern at similar magnitude but did not reach statistical significance, likely reflecting the low number of cases rather than absence of an effect. These findings align with previous evidence that Ch4/NbM is sensitive to early neurodegenerative change^9,17,56^, and extend it by showing within individuals and across diseases that this sensitivity may indeed be compartment-specific.

While cholinergic vulnerability is a shared feature of AD and Lewy body disorders (LBD), it appears to be regionally selective, dependent on the underlying pathology and disease stage, and strongly influenced by the measuring methods used. In AD, the most consistent decline involve the posterior, cortically projecting Ch4/NbM system, whereas the hippocampal-projecting Ch1–Ch2 regions are comparatively preserved^7,8,57^. In relatively pure AD, Ch4 degeneration is closely associated with tau pathology^58^. In LBD, post-mortem, neurochemical, PET, and MRI evidence most consistently implicates Ch4 and its cortical projections, with more pronounced degeneration and cholinergic dysfunction in cognitively impaired patients^12,16,56,59,60^. Ch1–Ch2 loss has been demonstrated in selected DLB cohorts and may affect their hippocampal projections, but it is not established as a general feature of early or cognitively normal PD and may emerge mainly in advanced Lewy body dementia patients^15,16^. Longitudinal MRI studies strengthen the temporal interpretation by showing that smaller Ch4 volume and impaired Ch4 projections microstructural integrity can predict subsequent cognitive decline^56,59,60^. Co-pathology is also likely to be a major determinant of BF vulnerability with mixed AD/LBD cases show particularly severe posterior BF atrophy, suggesting additive or interacting effects of tau and α-synuclein pathology^58,61^.

Interestingly, one of the strongest estimated effects we found was not in a well validated cholinergic disorder but in MS/demyelinating disease. While the hallmark of MS is demyelinating white matter lesions in the CNS^62^, degenerative processes in deep grey matter with global reduction of neuronal density and the presence of axonal damage have been shown^63^, including in the BF^64^. Furthermore, while BF projections are composed mainly of thin unmyelinated axons^18,54^, the effects of MS pathology are not confined to myelinated tracts, and are also demonstrated in unmyelinated fibres^65^. Previous evidence for retrograde axonal degeneration-induced BF damage^66^ points to a possible pathological mechanism in this disorder. Furthermore, reduced hippocampal synthesis of acetylcholine was found in both demyelinated and non-demyelinated hippocampi in MS^67^, aligning with our findings of reduced integrity of the hippocampal-innervating Ch1-3 in these patients. No psychiatric disorder showed structural BF deficits, including schizophrenia, which is characterized by cholinergic dysfunction and dysregulation. However, the strongest evidence in schizophrenia has been pointing to abnormal post-synaptic reduction of muscarinic and nicotinic cholinergic receptor availability in cortical, hippocampal and striatal circuits, rather than direct selective BF damage^20,68,69^. While a previous replicated structural MRI study identified lower BF volume in schizophrenia mainly in the medial septum, this difference was attenuated after adjustment for global structural brain measures, arguing against a selective BF vulnerability^70^.

Here, negative Ch4 deviations were also observed in participants scanned a median of four to five years before dementia or PD diangosis, regardless of pre-diagnosis cutoff definition. Structural changes in Ch4 were previously reported in the years before AD diagnosis^9,71^ and during early prodromal stages of LBD^56^. In addition, only pre-diagnostic dementia, but not PD, was associated with Ch1-3 deficits, therefore contributing disease- and compartment-specific evidence supporting selective vulnerability of the BF nuclei across the two disorders. Furthermore, the definition of pre-diagnostic we used in the current study, classified as receiving a diagnosis of dementia or PD more than 12 months after imaging, warrants reflection. This approach enabled the use of incident cases in this large population-based cohort, but it introduces potential look-ahead bias because group membership was determined retrospectively using future diagnostic information. These pre-diagnostic participants likely spanned a range of underlying disease stages, from truly preclinical pathology to unrecognized prodromal manifestations at the time of imaging.

Baseline Ch4 deviation was further associated with incident dementia and PD diagnosis, and did so independently of demographic and lifestyle factors, polygenic liability, and in some cases from other disease-related brain metrics. The Ch1-3 compartment, by contrast, carried no independent prospective signal and its univariable association with incident dementia was generally absorbed with Ch4 in the model. This asymmetry between the two compartments can be viewed as a prospective counterpart of the manifest-disease dissociation, and it was robust across the model specification ladder. Additionally, the Ch4 dementia association was not constant over the follow-up period but was more pronounced closer to imaging. An association strongest in the years closer to the scan and weakening thereafter could reflect pathology already present at imaging and becoming manifest as it approaches clinical threshold, rather than a more stable trait forecasting disease many years ahead. Therefore, such a marker might be more suitable for identification of people closer to the threshold of clinical disease over long-term risk stratification in mid-life. However, because these analyses are observational and rest, especially for the adjusted models, on modest event counts, they should be regarded as a population-scale proof-of-concept, rather than as definitive evidence of stand-alone prospective screening tools. Future validation will require large longitudinal imaging cohorts to determine whether the normative deviation scores improve risk stratification and pre-diagnostic classification beyond established criteria.

Lastly, we found that genetic liability to these disorders contributed little to Ch4 or Ch1-3 integrity, with very small and importantly, non-specific, associations. This is in accordance with previous works showing weak associations between polygenic risk scores and brain structure in UKB^72,73^. Given the size of the current sample, these significant results and tight intervals around near-zero effects are evidence of very weak relationships and argue against a substantially heritable common-variant basis for BF vulnerability. The finding does not exclude rare-variant or oligogenic contributions, nor gene–environment interactions, which warrants separate specific investigation. It therefore leaves open where the explanatory weight for BF vulnerability itself is more likely to lie, including age-related, environmental, and modifiable factors, which future work will need to examine directly.

Several limitations of the current work should be acknowledged. First, the data and subsequent analyses of the current study are observational and mostly cross-sectional and support no direct causal inference. It should also be noted that while in vivo “cholinergic” basal forebrain volumetry is often interpreted as though it were a cholinergic-specific marker, the volumetric deviation should be better framed as a more general structural vulnerability of this region. This is mainly due to the fact the BF contains not only cholinergic cells, but also other cell types such as GABAergic and glutamatergic^74^, a separation which MRI resolution cannot address. Furthermore, a significant proportion of the GM compartment in the brain has been suggested to be taken up by axonal and dendritic projections, and not primarily cell bodies^75^. Nevertheless, the higher vulnerability of Ch4 demonstrated in this study as well as previous works, may pinpoint to a higher vulnerability of cholinergic neurons specifically, given that over 90% of the Ch4 neurons have been proposed to be cholinergic, while this ratio appears much lower in the cell groups within Ch1-3^3,51^. Additionally, the disease groups defined from health records in this study are heterogeneous and, for some of the conditions modest in size. Further, to infer disease-related specificity, we defined cases as diagnostically mutually-exclusive, while in real-life co-morbidity among these conditions commonly takes place. This is important given the well-known selection bias of the UKB, which may not optimally reflect the true distribution in the general population and limits direct generalization to real-world clinical settings^76^. Accordingly, the present results should be interpreted as population-based proof-of-concept evidence rather than direct evidence of clinical utility.

### Conclusions

The human basal forebrain is not a single, uniformly vulnerable system: its cholinergic compartments dissociate across every axis examined here. The Ch4/NbM ages on its own trajectory from mid-life, is disproportionately affected across neurological disease, shows deviation years before diagnosis, and independently predicts incident dementia and PD, largely independent of polygenic risk for the conditions studied. The hippocampus-projecting Ch1-3/septal-diagonal band compartment, by contrast, is affected in fewer conditions, and where it does show an association, as in dementia, this carries no independent prognostic value once Ch4 is accounted for. These findings argue for treating the basal forebrain as a set of anatomically and functionally distinct compartments rather than a single structure and identify Ch4/NbM deviation from population norms as an anatomically specific, pre-diagnostic marker of neurodegenerative vulnerability. More generally, the normative modelling framework offers a scalable approach for quantifying selective regional vulnerability as an age-related process, and a foundation for investigating mechanisms of resistance, resilience, and risk in normal ageing and neurodegenerative disease.

### Data availability

UK Biobank data are available for approved researchers through a procedure described at https://www.ukbiobank.ac.uk/enable-your-research.

### Code availability

Code used for this publication will be made available via GitHub upon acceptance.

## Acknowledgments

For the purpose of open access, the authors have applied a CC BY public copyright license to any Author Accepted Manuscript version arising from this submission.

## Funding

This research was supported by the NIHR Oxford Health Biomedical Research Centre (NIHR203316). TE is supported by the NIHR Oxford Biomedical Research Centre NIHR203311. JCK acknowledges support from the NIHR Oxford Health Clinical Research Facility, and the NIHR Oxford Biomedical Research Centre. The Centre for Integrative Neuroimaging was supported by core funding from the Wellcome Trust (203139/Z/16/Z and 203139/A/16/Z).

## Author contribution

T.E: Conceptualization, Methodology, Investigation, Formal analysis, Writing - Original Draft, Writing - Review & Editing; M.H: Funding acquisition, Project administration; J.K: Conceptualization, Methodology, Writing - Review & Editing, Supervision, Project administration

## Conflict of interest statement

The authors declare no competing financial or non-financial interests or any conflict of interest.

## Consent statement

All participants provided written informed consent, and the study was approved by the National Information Governance Board for Health and Social Care and the National Health Service North-West Multicentre Research Ethics Committee (21/NW/0157, see http://www.ukbiobank.ac.uk/ethics/).

## Notes

### Competing Interest Statement

The authors have declared no competing interest.

### Author Declarations

Written informed consent of all participants was obtained by UK Biobank. Permission to use the UKB Resource was obtained as part of Data Access Application 95506. All participants provided written informed consent, and the study was approved by the National Information Governance Board for Health and Social Care and the National Health Service North-West Multicentre Research Ethics Committee (21/NW/0157, see http://www.ukbiobank.ac.uk/ethics/).

