## Supplementary Information for "Selective vulnerability of human basal forebrain subsystems in ageing and neuropsychiatric disorders"

**Supplementary Table 1.** Diagnostic codes for exclusion from the healthy reference sample derived from UK Biobank data **category 1712**.

| **Diagnosis** | **Code** |
| --- | --- |
| Ischemic/ hypertensive heart disease | I11, I12, I13, I15, I20, I21, I22, I23, I24, I25, I26, I27, I46, I50 |
| Stroke/  cerebrovascular disease | I60, I61, I62, I63, I64, I65, I66, I67, I68, I69 |
| COPD | J43, J44 |
| Chronic kidney disease (CKD) | N18 |
| All-cause dementia | F00, F01, F02, F03, F04, G30 |
| Parkinson’s disease | G20 |
| Other Parkinsonism | G21, G22, G23, G24, G25 |
| Multiple sclerosis and demyelination diseases of CNS | G35, G36, G37 |
| Diabetes | E11 |
| Cirrhosis | K70, K71, K72, K74 |
| Schizophrenia | F20, F21, F22, F23, F24, F25 |
| Depression | F33 |
| Bipolar disorder | F31 |
| Other neurodegenerative disease | G10, G31, G32 |
| Cancer | Cancer diagnosis was extracted from **Data-Field 40005** |

**Supplementary Table 2.** Model performance for the different distribution families that converged in each metric (presenting top 5 BIC for each side)

| R CH4 GM Vol (n=43,620) | | | R CH1-3 GM Vol (n=43,591) | | | L CH4 GM Vol (n=43,616) | | | L CH1-3 GM Vol (n=43,586) | | |
| --- | --- | --- | --- | --- | --- | --- | --- | --- | --- | --- | --- |
| family | BIC | converged | family | BIC | converged | family | BIC | converged | family | BIC | converged |
| JSU | -132750 | TRUE | JSU | -110677 | TRUE | JSU | -145695 | TRUE | JSU | -112073 | TRUE |
| BCT | -132703 | TRUE | BCTo | -110647 | TRUE | JSUo | -145670 | FALSE | BCTo | -112035 | TRUE |
| BCTo | -132700 | TRUE | BCT | -110646 | TRUE | BCTo | -145658 | TRUE | BCT | -112035 | TRUE |
| BCPE | -132682 | TRUE | BCPEo | -110615 | TRUE | BCT | -145658 | TRUE | BCPEo | -112002 | TRUE |
| BCPEo | -132679 | TRUE | BCPE | -110614 | TRUE | BCPEo | -145590 | TRUE | BCPE | -112002 | TRUE |

**Supplementary Table 3.** Models variables’ UK Biobank data fields.

| **Variable** | **Data field** |
| --- | --- |
| Age | Derived from 33 and 53 |
| Sex | 31 |
| Townsend deprivation index | 189 |
| Educational attainment | Derived from 6138 |
| Smoking status | Derived from 20116 |
| Alcohol-intake frequency | 1558 |
| Body mass index (BMI) | 21001 |
| Polygenic risk scores | G35, G36, G37 |
| Hippocampal volume | Derived from 25886, 25887 |
| Cortical grey matter volume | 25002 |
| Substantia nigra magnetic susceptibility | Derived from 24481, 24482 |

**Supplementary Table 4.** Model performance for the different distribution families that converged in the disease-specific brain metrics (presenting top 5 BIC for each side)

| Cortical GM volume | | | B Hippocampus GM volume | | | Bilateral SN susceptiblity | | |
| --- | --- | --- | --- | --- | --- | --- | --- | --- |
| family | BIC | converged | family | BIC | converged | family | BIC | converged |
| NO | 1001550 | TRUE | ST4 | 684838.8 | TRUE | JSU | 326447.6 | TRUE |
| JSU | 1001552 | TRUE | ST3 | 684884.2 | TRUE | SEP4 | 326449.1 | TRUE |
| ST4 | 1001553 | TRUE | JSU | 684885.6 | TRUE | SHASH | 326462.8 | TRUE |
| BCTo | 1001569 | TRUE | BCTo | 684892.8 | TRUE | ST3 | 326474.8 | TRUE |
| SHASHo | 1001570 | TRUE | BCT | 684896.8 | TRUE | BCT | 326476.8 | TRUE |

**Supplementary Table 5.** Cox models sample sizes.

| **Outcome** | **Model** | **N** | **Events/cases** |
| --- | --- | --- | --- |
| All-cause dementia | Minimal | 53,477 | 129 |
| All-cause dementia | Minimal + general covariates | 51,208 | 115 |
| All-cause dementia | Minimal + general covariates + PRS | 49,733 | 109 |
| All-cause dementia | Minimal + general covariates + brain metrics | 51,190 | 115 |
| All-cause dementia | Minimal + general covariates + PRS + brain metrics | 49,715 | 109 |
| Parkinson's disease | Minimal | 53,477 | 76 |
| Parkinson's disease | Minimal + general covariates | 51,208 | 70 |
| Parkinson's disease | Minimal + general covariates + PRS | 49,733 | 66 |
| Parkinson's disease | Minimal + general covariates + brain metrics | 46,494 | 57 |
| Parkinson's disease | Minimal + general covariates + PRS + brain metrics | 45,137 | 53 |

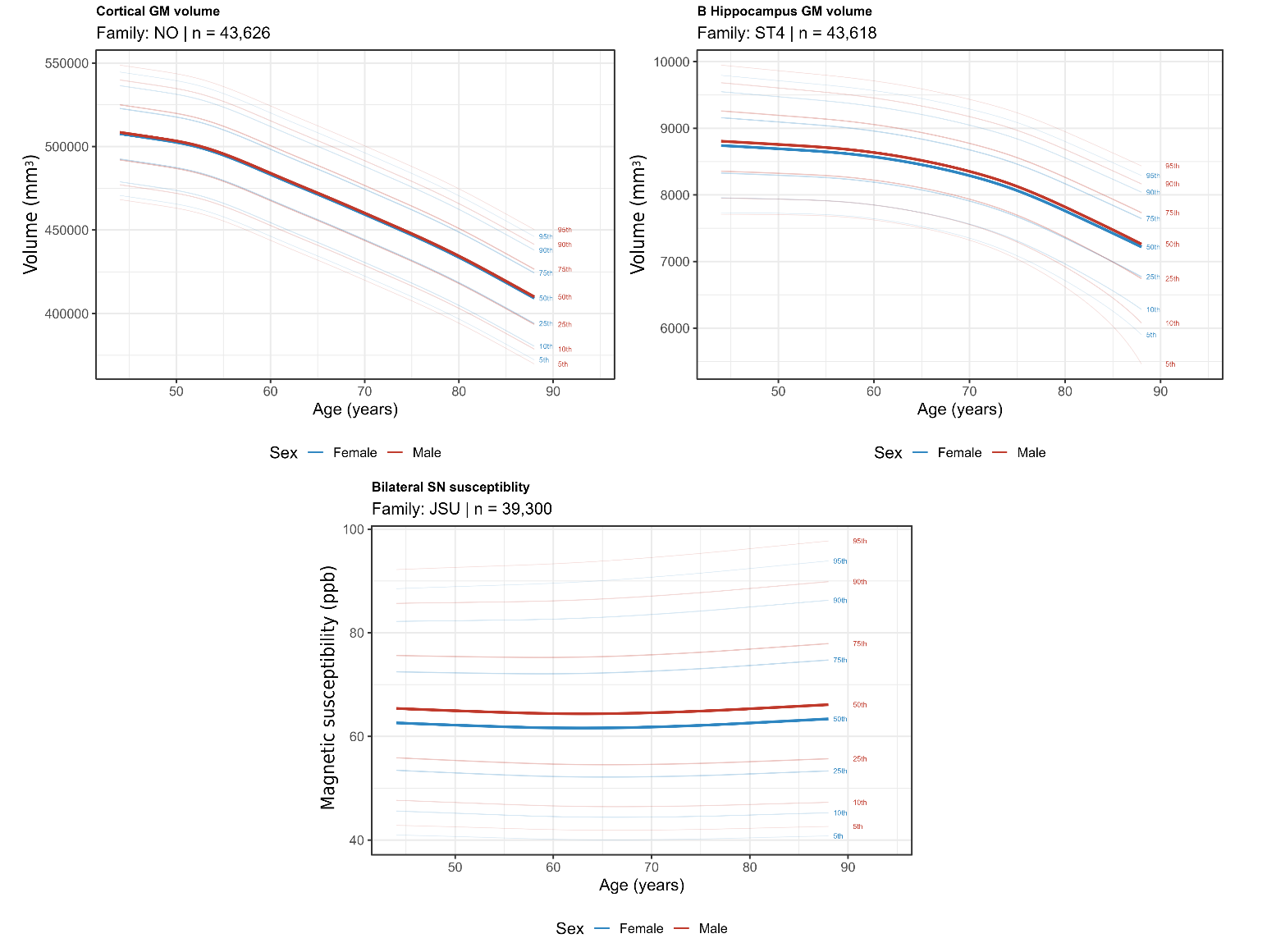

**Supplementary Figure 1. Normative trajectories from mid to late life of the disease-specific brain metrics.** Population centile curves for cortical GM volume (top left), bilateral hippocampal GM volume (top right) and bilateral substantia nigra magnetic susceptibility (bottom), from generalised additive models for location, scale and shape (GAMLSS) fitted on a healthy reference sample of UK Biobank participants aged 45–87 years. Curves show the 5th, 10th, 25th, 50th, 75th, 90th and 95th centiles as a function of age, modelled separately by sex (blue, female; red, male). Selected distribution families and the available healthy reference sample for each metric are presented. B = bilateral; GM = grey matter; ppb = parts per billion; SN = substantia nigra.

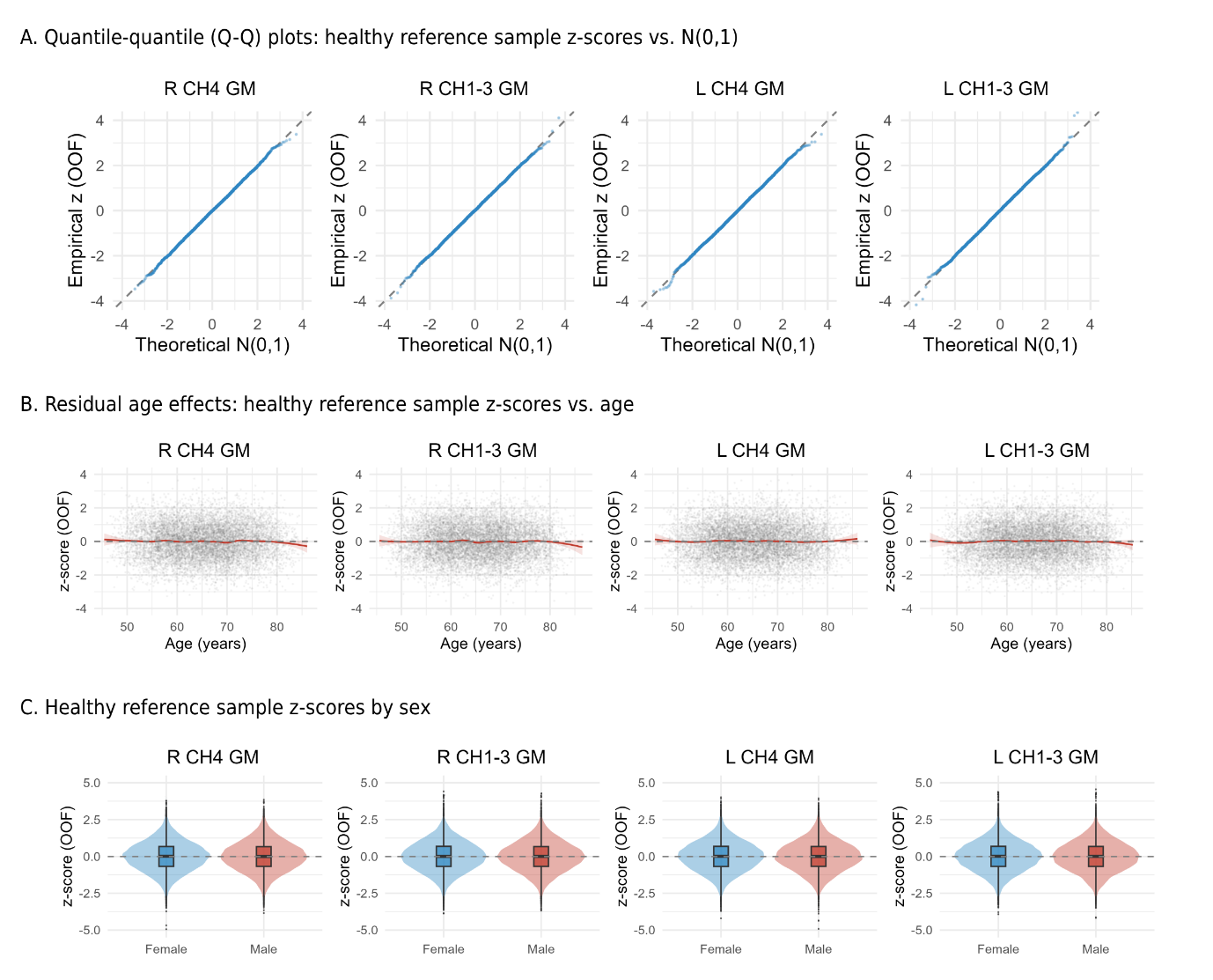

**Supplementary Figure 2. Quality control metrics of the normative models.** Quality-control diagnostics for the generalised additive models for location, scale and shape (GAMLSS) used to derive normative deviation (z) scores of the basal forebrain grey-matter volume, shown for each hemisphere and compartment (R/L nucleus basalis, Ch4; R/L septal/diagonal-band, Ch1–3). All diagnostics are based on the out-of-fold (OOF) z-scores in the healthy reference sample, so that each participant's deviation is computed from a model not fitted on that participant. **(A)** Quantile–quantile plots of the empirical OOF z-scores against the theoretical standard normal distribution; points falling on the identity line (dashed) indicate that the modelled distribution is well calibrated, with only minor departures in the extreme tails. **(B)** OOF z-scores as a function of age; the flat trend (red, with 95% confidence band; grey points are individual participants) indicates that age has been adequately modelled and that no systematic age-related bias remains in the deviation scores across the sampled range (45–87 years). **(C)** Distribution of OOF z-scores by sex (split violin plots with overlaid box plots; blue, female; red, male), centred on ~0 with comparable spread in both sexes, indicating the models are well calibrated across sex. Together these diagnostics confirm that the normative deviation scores are approximately standard-normal, free of residual age or sex bias, and therefore comparable across region, hemisphere, age and sex in the downstream analyses. GM = grey matter; L = left; OOF = out-of-fold; R = right

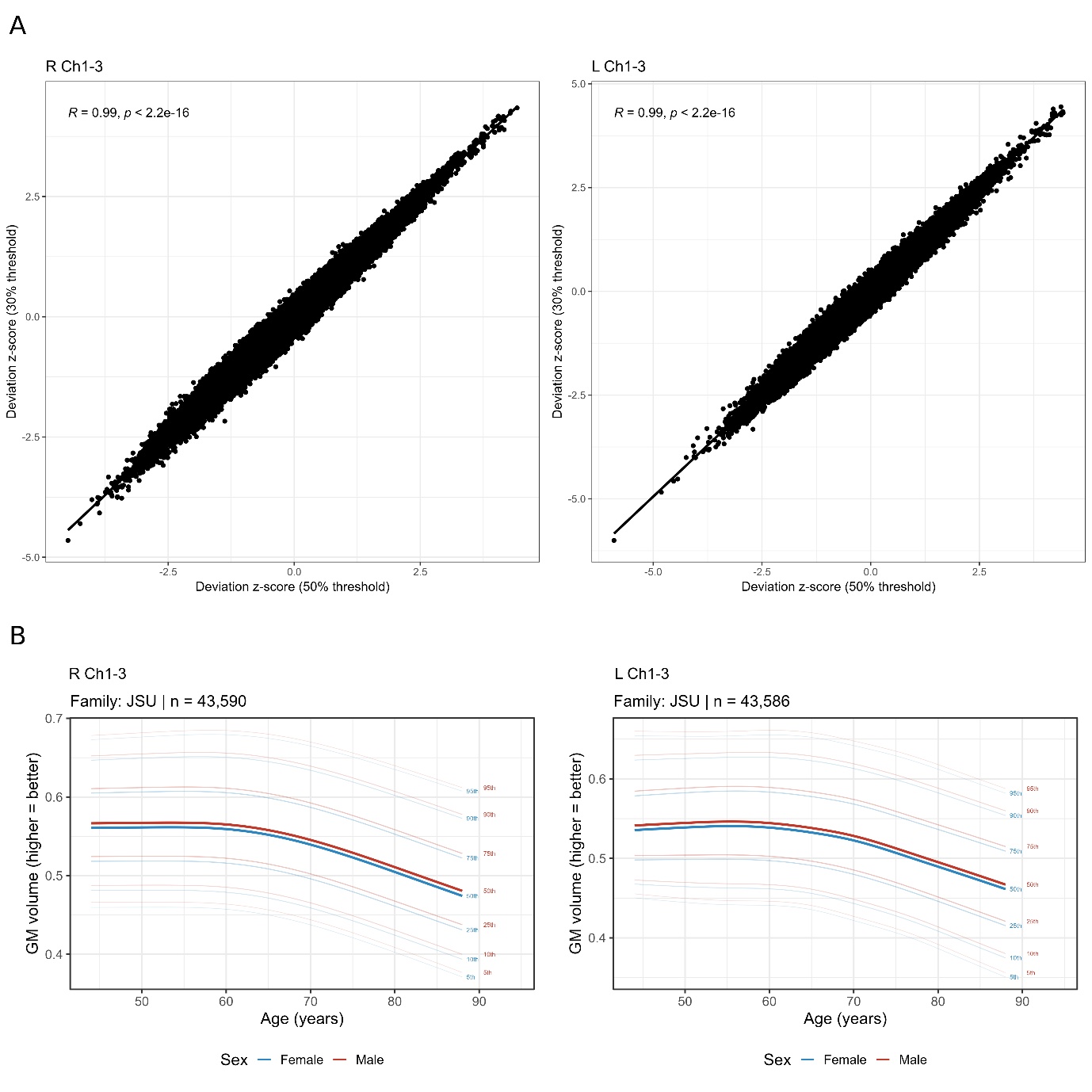

**Supplementary Figure 3. Normative modelling of the Ch1-3 with a 30% threshold.** (A) Pearson correlations between the deviation z-scores derived from the 50%- and 30%-threshold models for the right and left Ch1-3. (B) Population centile curves of grey matter (GM) volumes for the right and left Ch1-3 from the generalised additive models for location, scale and shape (GAMLSS) fitted on a healthy reference sample of UK Biobank participants aged 45–87 years. Curves show the 5th, 10th, 25th, 50th, 75th, 90th and 95th centiles as a function of age, modelled separately by sex (blue, female; red, male). Selected distribution families and the available healthy reference sample for each metric are presented.

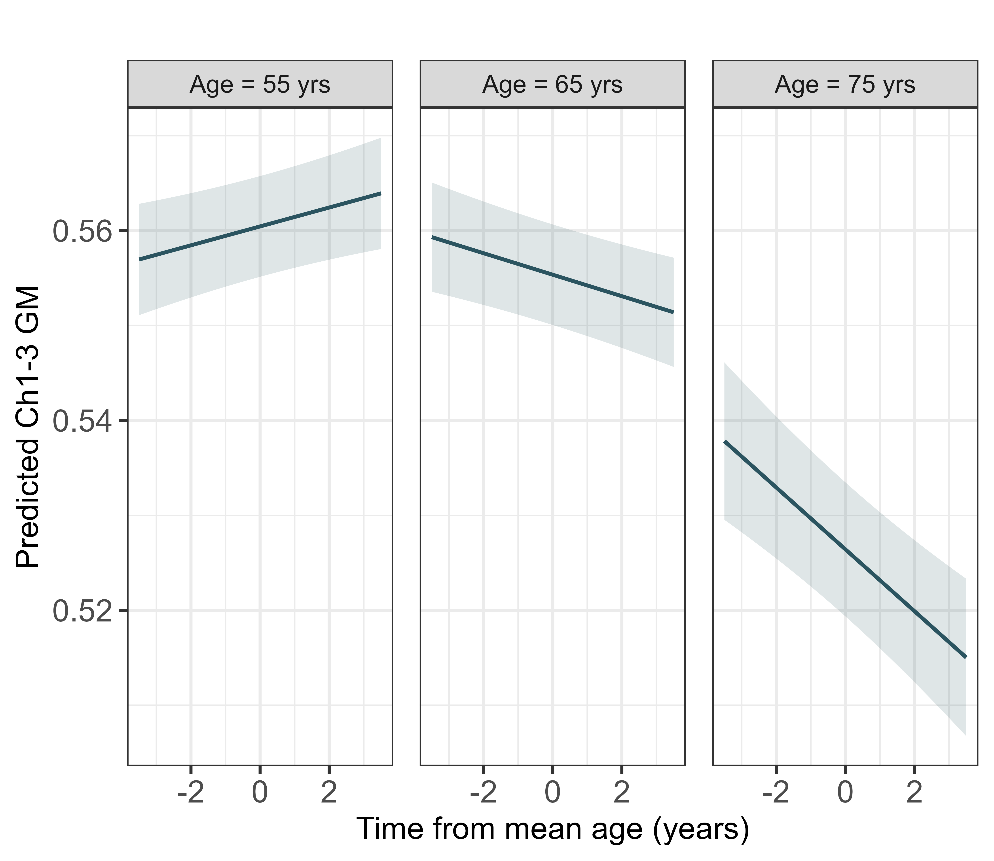

**Supplementary Figure 4. Ch1-3 GM volume change over time with a 30% threshold.** Age-related changes over time in grey matter volume of the Ch1-3 (based on a 30% threshold) using generalized additive mixed models (GAMM) performed on a subset of the healthy reference sample with two time points of imaging data (n=2,712). The model-based predicted grey matter volume changes are presented for three different ages (55, 65, 75 years) showing that Ch1-3 decline from the seventh decade.

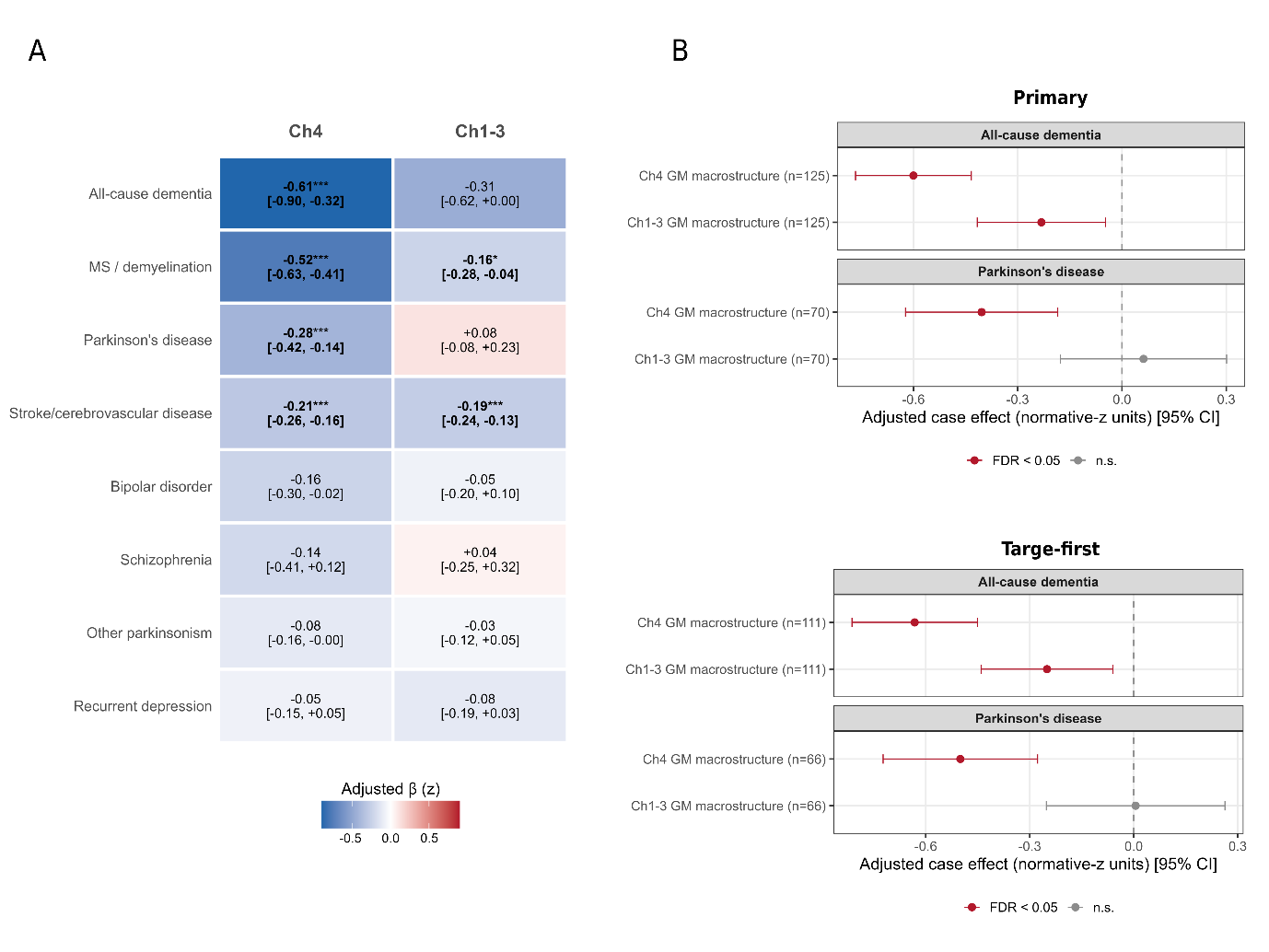

**Supplementary Figure 5. Matched case-control analysis of manifest and pre-diagnostic cases.** (A) Adjusted case–control differences in normative deviation (β, in age- and sex-conditioned z-units; 95% CI) for the nucleus basalis (Ch4) and the septal/diagonal-band compartment (Ch1–3) across eight conditions, from covariate-matched (case-control ratio 1:5) comparisons (Methods). Negative values (blue) indicate lower-than-expected grey-matter volume for age and sex; colour encodes effect magnitude and direction, and asterisks with bold type denote associations surviving FDR-correction (* *p*FDR *< 0.05, ** p*FDR *< 0.01,* *** *p*FDR < 0.001). (B) Adjusted case–control differences in deviation (normative-z units, 95% CI) between participants who received a first diagnosis of all-cause dementia or Parkinson's disease at least 12 months after imaging and a disease-free reference sample, estimated from the covariate-matched (case-control ratio 1:5) comparisons, for the primary and target-first definitions.

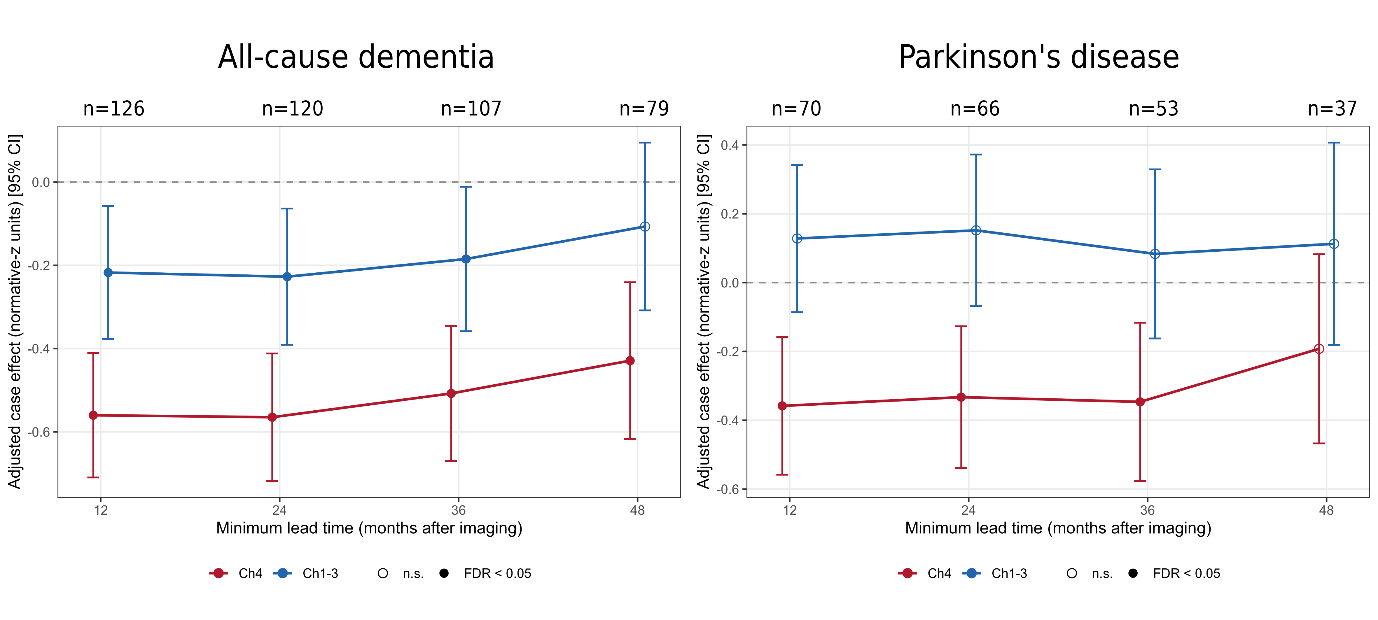

**Supplementary Figure 6. The pre-diagnostic Ch4 deficit is robust to the cut-off definition.** Adjusted case effect (normative-z units, ±1 SE) the Ch4 (red) and Ch1–3 (blue) grey-matter deviation in future cases of all-cause dementia (left) and Parkinson's disease (right), as a function of the minimum required interval between imaging and future diagnosis (12, 24, 36, 48 months). The horizontal axis represents progressive removal of the cases nearest to diagnosis. At each cut-off, the model, covariates, and healthy reference sample are identical to the primary pre-diagnostic analysis. Filled points indicate associations surviving FDR correction within each cut-off while open points do not. Case numbers at each cut-off are shown above the panel. Error bars denote 95% confidence interval.

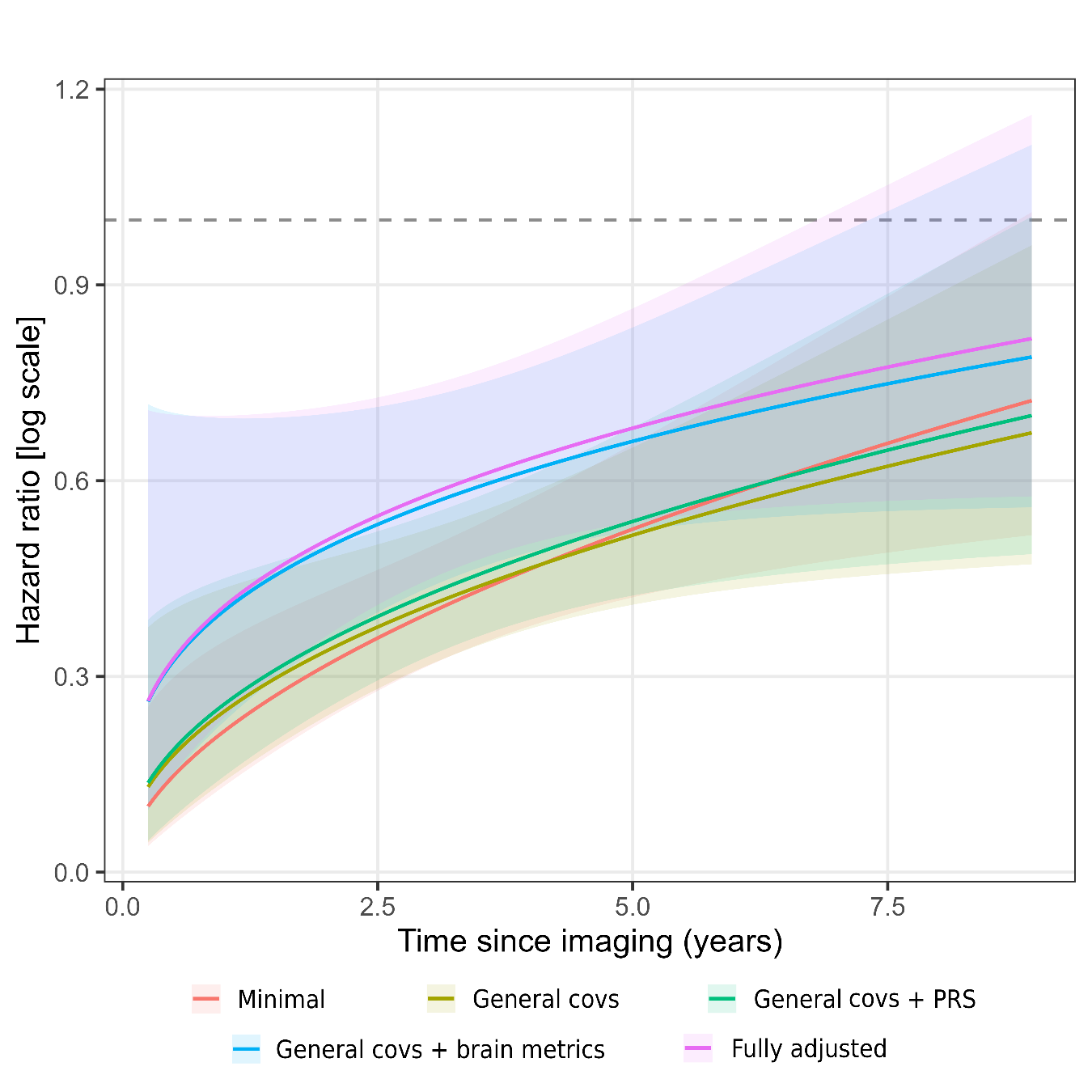

**Supplementary Figure 7. Continuous time-varying hazard ratio for Ch4 and incident dementia.** Reconstructed hazard-ratio function HR(t) = exp(β₀ + β₁·log t) for Ch4 grey-matter deviation and incident all-cause dementia, from the joint Cox models in which the Ch4 coefficient was allowed to vary continuously with time via a time-transform (tt) term (Methods). Each line is one of the models and covariate set, and shaded bands are 95% confidence intervals. The hazard ratio is per +1 normative-z (higher = greater-than-expected volume). The dashed line marks HR = 1 (no association). Across all model sets the Ch4 hazard ratio is furthest below unity shortly after imaging and rises toward 1 with increasing follow-up, indicating an association concentrated closer to the scan that attenuates over time; the coefficient remains below unity across the range supported by the data. Confidence bands widen at the earliest and latest follow-up times, where log-time is extreme and events are sparse. Dashed line represents HR of 1 (no association). Covs = covariates; PRS = polygenic risk score
